# The Impact of Transcutaneous Spinal Cord Stimulation on Autonomic Regulation after Spinal Cord Injury: A randomized crossover trial

**DOI:** 10.1101/2023.07.18.23292676

**Authors:** Ryan Solinsky, Kathryn Burns, Christopher Tuthill, Jason W. Hamner, J. Andrew Taylor

## Abstract

**Importance:** Individuals with spinal cord injury (SCI) have significant autonomic nervous system dysfunction. However, despite recent findings postulated to support that spinal cord stimulation improves dynamic autonomic regulation, limited scope of previous testing means the true effects remain unknown.

**Objective:** To determine whether transcutaneous spinal cord stimulation improves dynamic autonomic regulation after SCI.

**Design:** Single-blinded, randomized crossover trial with matched cohorts.

**Setting:** Academic autonomic physiology laboratory.

**Participants:** Two pairs of well-matched individuals with and without high-thoracic, complete SCI.

**Interventions:** Sub-motor threshold transcutaneous spinal cord stimulation delivered at T10-T11 using 120Hz, 30Hz, and 30Hz with 5kHz carrier frequency at separate autonomic testing sessions.

**Main Outcomes and Measures:** Baseline autonomic regulation was characterized with tests of above injury level sympathoexcitation (Valsalva’s maneuver), sympathoinhibition (progressive doses of bolus intravenous phenylephrine), and below level sympathoexcitation (foot cold pressor test). At three subsequent visits, this testing battery was repeated with the addition of spinal cord stimulation at each frequency. Changes in autonomic regulation for each frequency were then analyzed relative to baseline testing for each individual and within matched cohorts.

**Results:** Uninjured controls demonstrated no autonomic deficits at baseline and had no changes with any frequency of stimulation. Contrasting this, and as expected, individuals with SCI had baseline autonomic dysfunction. In a frequency-dependent manner, spinal cord stimulation enhanced sympathoexcitatory responses, normalizing previously impaired Valsalva’s maneuvers. However, stimulation exacerbated already impaired sympathoinhibitory responses, resulting in significantly greater mean arterial pressure increases with the same phenylephrine doses compared to baseline. Impaired sympathoexcitatory response below the level of injury were also further exacerbated with spinal cord stimulation. At baseline, neither individual with SCI demonstrated autonomic dysreflexia with the noxious foot cold pressor test; the addition of stimulation led to a dysreflexic response in every trial, with greater relative hypertension and bradycardia indicating no improvement in autonomic regulation.

**Conclusions and Relevance:** Transcutaneous spinal cord stimulation does not improve autonomic regulation after SCI, and instead likely generates tonic, frequency-dependent sympathoexcitation which may lower the threshold for autonomic dysreflexia.

**Trial Registration:** Transcutaneous Spinal Cord Neuromodulation to Normalize Autonomic Phenotypes; NCT04858178. https://clinicaltrials.gov/ct2/show/NCT04858178

**Key Points:**

- Does electrical spinal cord stimulation, at any previously advocated stimulation frequency, improve regulation of the autonomic nervous system for individuals with spinal cord injuries?
- In this randomized crossover trial, transcutaneous spinal cord stimulation generated tonic, frequency-dependent sympathetic activation below the level of injury, without improved dynamic autonomic regulation.
- Tonic sympathetic activation below the level of injury could lower the threshold for potentially dangerous autonomic dysreflexia in individuals with spinal cord injury and future work should employ appropriate monitoring.

## Introduction

Individuals with spinal cord injury (SCI) commonly have blood pressure instability, manifesting as orthostatic hypotension (insufficient sympathetic activation through the injured cord to maintain adequate blood pressure)^1^ and/or autonomic dysreflexia (AD, potentially dangerous hypertension triggered by noxious stimuli below the level of injury, causing regional sympathoexcitation that cannot be counter-regulated).^2^ After SCI, descending regulatory brainstem signals are interrupted, decoupling important spinal cord sympathetic circuitry and compromising blood pressure regulation.^3^ To address this, multiple recent small, high-impact studies have utilized spinal neuromodulation to stabilize blood pressure during orthostatic challenge.^4-7^ Though demonstrating proof of principle, differing stimulation frequencies and limited physiologic monitoring/challenges provide a narrow view of autonomic nervous system responses. If regulation is improved, both sympathoexcitatory and sympathoinhibitory responses should be enhanced. Orthostatic challenge requires sympathoexcitation alone; hence, stimulation may simply induce a hypertensive response via local, unperceived noxious stimuli (AD).^8^ Given ongoing industry-sponsored clinical trials and the current limited understanding, a more comprehensive characterization of the autonomic regulatory effects of spinal cord stimulation is critically needed.

## Methods

Individuals 18-30 years of age with chronic high thoracic motor/sensory complete SCI (E-ISNCSCI exam^9)^ and matched uninjured controls were enrolled from February to November 2022 (Figure 1A). Beat-to-beat blood pressure, heart rate, respiration, and leg blood flow were recorded in our academic physiology laboratory. Data during 5-minutes of resting baseline were obtained, followed by Valsalva’s maneuvers, bolus intravenous phenylephrine, and cold pressor test. Valsalva’s maneuver, a sustained forced exhalation against resistance generates characteristic responses across four phases. Phase II, the initial fall in pressure due to intrathoracic pressure restricting venous return to the heart requires sympathetic activation to maintain pressure.^10^ Bolus intravenous phenylephrine (three progressive doses^11^ with a goal of >15mmHg increase in systolic pressure (SBP))^11^ is normally buffered by bradycardia and sympathetic inhibition. The cold pressor test (three minutes of the foot emersed in 0⁰C ice water) results in increased blood pressure from tachycardia and sympathoexcitation.^13^ These three tests characterize both sympathoexcitatory and sympathoinhibtory responses through different reflex pathways. For the last, a noxious stimulus below the injury level in SCI further may increase blood pressure due to AD, resulting in reflexive baroreflex-mediated bradycardia rather than tachycardia.

**Figure 1:**
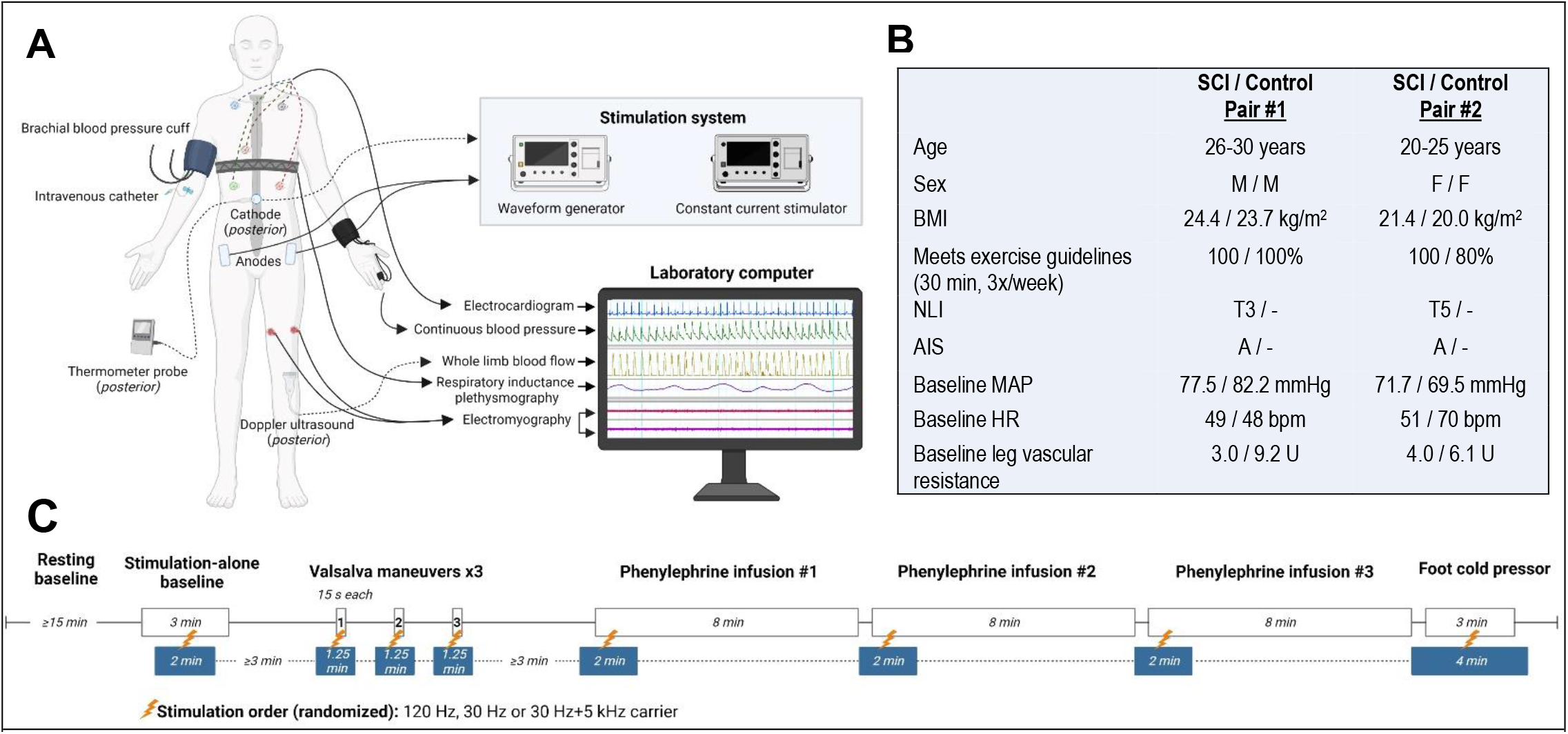
**A)** Participants were instrumented with five-lead electrocardiogram (Dinamap Dash 2000, GE) for continuous heart rate, respiratory inductance plethysmography (Respitrace, SensorMedics) for continuous breathing pattern, automated finger photoplethysmography (Finapres, Ohmeda Medical) for beat-to-beat arterial pressure in the left hand, and oscillometric cuff on the contralateral side (Dinamap Dash 2000, GE) for brachial arterial pressure and calibration with the finger cuff. Popliteal flow velocity was recorded from a 4-MHz Doppler ultrasound (Doppler-BoxX, DWL) at the left popliteal fossa for whole limb blood flow. An intravenous catheter was inserted in the right antecubital fossa by a trained medical provider. A 3.2cm circular electrode was placed at T10-T11 (acting as a cathode) and 5×9cm rectangular electrodes were placed at the bilateral iliac crests (acting as anodes). A thermometer probe was positioned at the cathode to ensure temperatures ≤38.5°C were maintained. The electrodes were connected to the stimulation system, consisting of a biphasic constant current stimulator (DS8R, Digitimer) triggered by a waveform generator (DG1022, RIGOL). Following a baseline testing session, the order of frequencies was randomized using a sequence generator for subsequent study sessions separated by at least seven days to account for any cumulative effects. All participants were blinded to the stimulation frequency. Surface electromyography (Dinamap Dash 2000, GE) was recorded in the left leg during autonomic testing to ensure sub-motor threshold stimulation. All data were digitally collected and stored on a computer at a sampling rate of 1,000Hz. This study was approved by the Mass General Brigham IRB and registered in ClinicalTrials.gov (NCT04858178). **B)** Demographics and resting baseline characteristics for two matched SCI and uninjured control pairs. Expectedly, lower extremity vascular resistance at baseline is decreased after SCI compared to controls (p=0.003). **C)** Study session outline consisting of stimulation alone, tests of above level sympathoexcitation (three Valsalva’s maneuvers), sympathoinhibition (three progressive doses of intravenous phenylephrine), and below level sympathoexcitation (foot cold pressor test) with and without transcutaneous spinal cord stimulation. Abbreviations. BMI= Body Mass Index, NLI= Neurological Level of Injury^10^, AIS= ASIA Impairment Scale^10^, MAP= Mean Arterial Pressure, HR= Heart Rate

Testing was repeated across four sessions with and without the addition of transcutaneous spinal cord stimulation at 120Hz,^4^ 30Hz,^5^ or 30Hz with 5kHz carrier frequency^6,7^ over T10-T11 (Figure 1). Amplitude was set at 80% of motor activation thresholds to prevent contraction-related hemodynamic contributions. One frequency was tested per session in a randomized order set by a sequence generator with participants blinded. Over four testing sessions for each individual, a rich dataset of 92 autonomic tests was generated to evaluate autonomic regulatory effects of stimulation.

For Valsalva’s maneuver, absolute decrease in mean arterial pressure (MAP) during Phase II, Valsalva’s ratio, and pressure recovery time^14^ were calculated. For bolus phenylephrine, absolute changes in MAP, heart rate (HR), and lower extremity vascular resistance were averaged across the three progressive doses of phenylephrine. As an equal comparator, the 75μg dose for those with SCI and matched controls were also similarly compared. For foot cold pressor, peak ΔMAP and ΔHR were calculated. Paired t-tests were completed between matched pairs, with stimulation at all frequencies grouped when data trends were homogenous. Results are reported as mean ±SE, with P<0.05 treated as significant.

## Results

Four individuals (Figure 1) completed all study sessions. With stimulation alone, MAP, HR, and leg vascular resistance were unchanged (both in aggregate and per frequency, eFigure 1).

At baseline, phase II pressure stabilization of Valsalva’s maneuver was present in controls and reliably absent in both individuals with SCI across all trials. MAP decrease during phase II was significantly greater at baseline for individuals with SCI than controls (−20.3±5.5 vs +4.9±2.4mmHg, p=0.01). Stimulation variably restored phase II pressure stabilization in a frequency dependent manner (Figure 2), with no such changes for matched controls. 120Hz stimulation normalized blood pressure in those with SCI compared to controls (mean decrease of 1.1±2.0mmHg, p=0.20) whereas 30Hz and 30Hz+5kHz stimulation were ineffective or less effective. Although pressure recovery time was not significantly different at baseline between those with SCI and controls (8.7±3.1sec for SCI vs 2.2±0.8sec for controls, p=0.09), stimulation at all frequencies lengthened pressure recovery time for individuals with SCI. Valsalva’s ratios were not different at baseline and stimulation had no effect.

**Figure 2:**
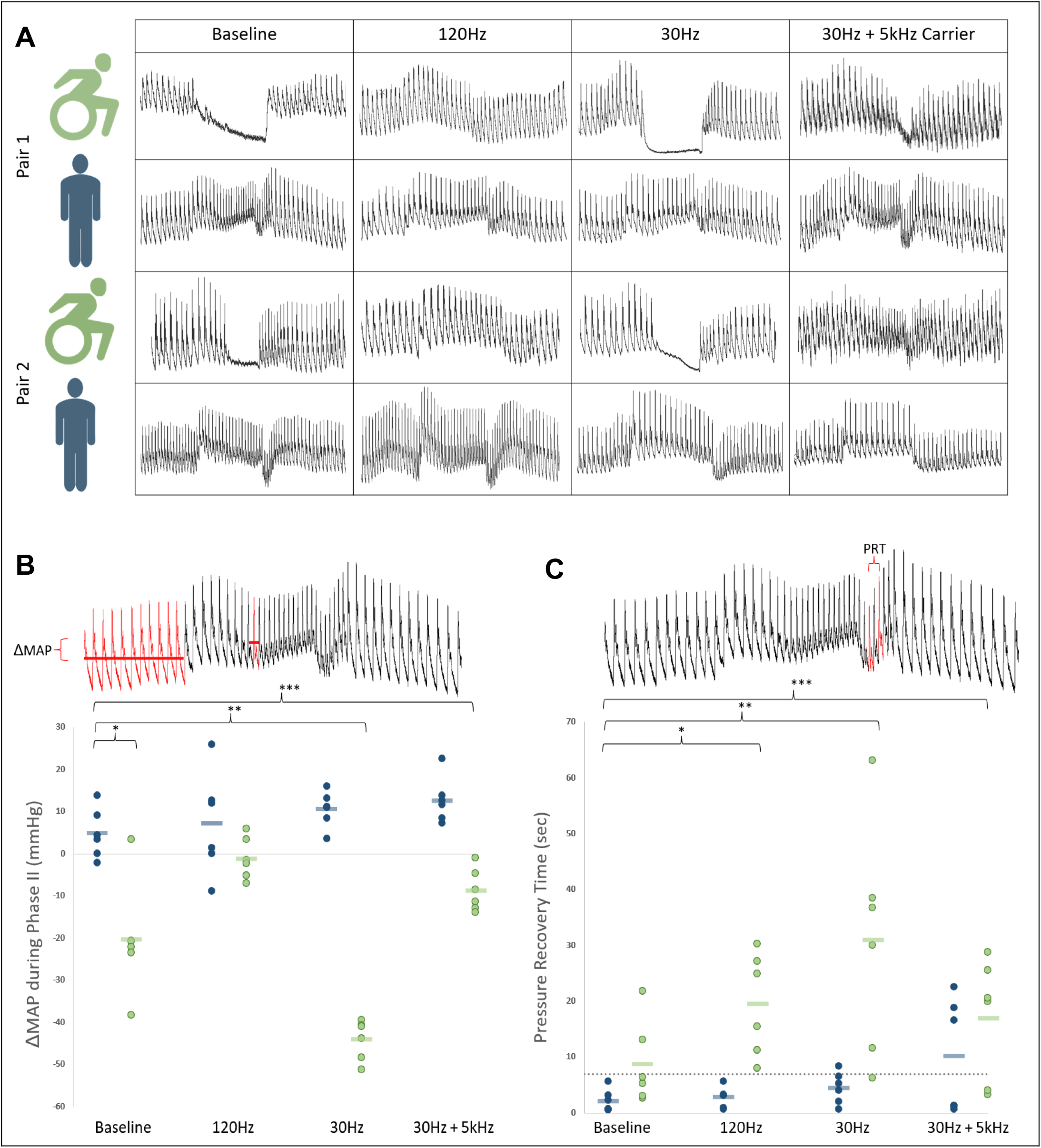
**A)** Valsalva’s maneuver at a goal sustained pressure of 30mmHg for 15 seconds. Representational traces of beat-to-beat blood pressure at baseline and with each stimulation frequency for all individuals. Of note, both individuals with SCI at baseline lack phase II stabilization of blood pressure which was subsequently restored with 120Hz. **B)** MAP changes from baseline to nadir of phase II of Valsalva’s maneuver. Group means represented as 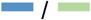 **C)** Pressure recovery time from nadir MAP in phase III to return to baseline MAP in phase IV. Dashed line represents upper limits of normal. In SCI, this time is significantly lengthened at baseline – an effect which is accentuated with transcutaneous spinal cord stimulation at all frequencies (120Hz, p=0.005; 30Hz, p=0.02; 30Hz+5kHz, p=0.03). Abbreviations. MAP= Mean Arterial Pressure, PRT= Pressure Recovery Time, *,**,***= statistically significant

Bolus phenylephrine tended to produce greater ΔMAP and relative resistance in SCI than control, though this did not reach significance (p=0.21 and p=0.16 respectively). Across all settings (n=18 trials) compared to baseline (n=6 trials), stimulation significantly phenylephrine-induced ΔMAP in SCI (27.0±2.4mmHg vs 6.8±1.1mmHg, p<0.001) but not in control (4.5±1.2mmHg vs 6.3±0.6mmHg, p=0.36). Similarly, vascular resistance was significantly increased with stimulation in SCI (+0.68±0.03U vs +1.6±0.19U, p=0.001), but not in controls (+0.12±0.31U vs +0.01±0.22U, p=0.79).

The foot cold pressor resulted in small MAP increases and a slight tachycardia that were unaltered by stimulation in controls (Figures 3C/D). In contrast, foot cold pressor resulted in small MAP increases and a modest bradycardia that were enhanced by stimulation in those with SCI. Across all stimulation frequencies, SBP increased on average by 27.7±4.5mmHg accompanied by a bradycardia of 6.3±1.3 beats. The greatest effect was seen at 120Hz (Figure 3). Hence, without stimulation in those with SCI, the foot cold pressor evidenced a modest noxious response that did reach threshold for AD, but stimulation augmented this such that every trial of the cold pressor test resulted in AD.

**Figure 3:**
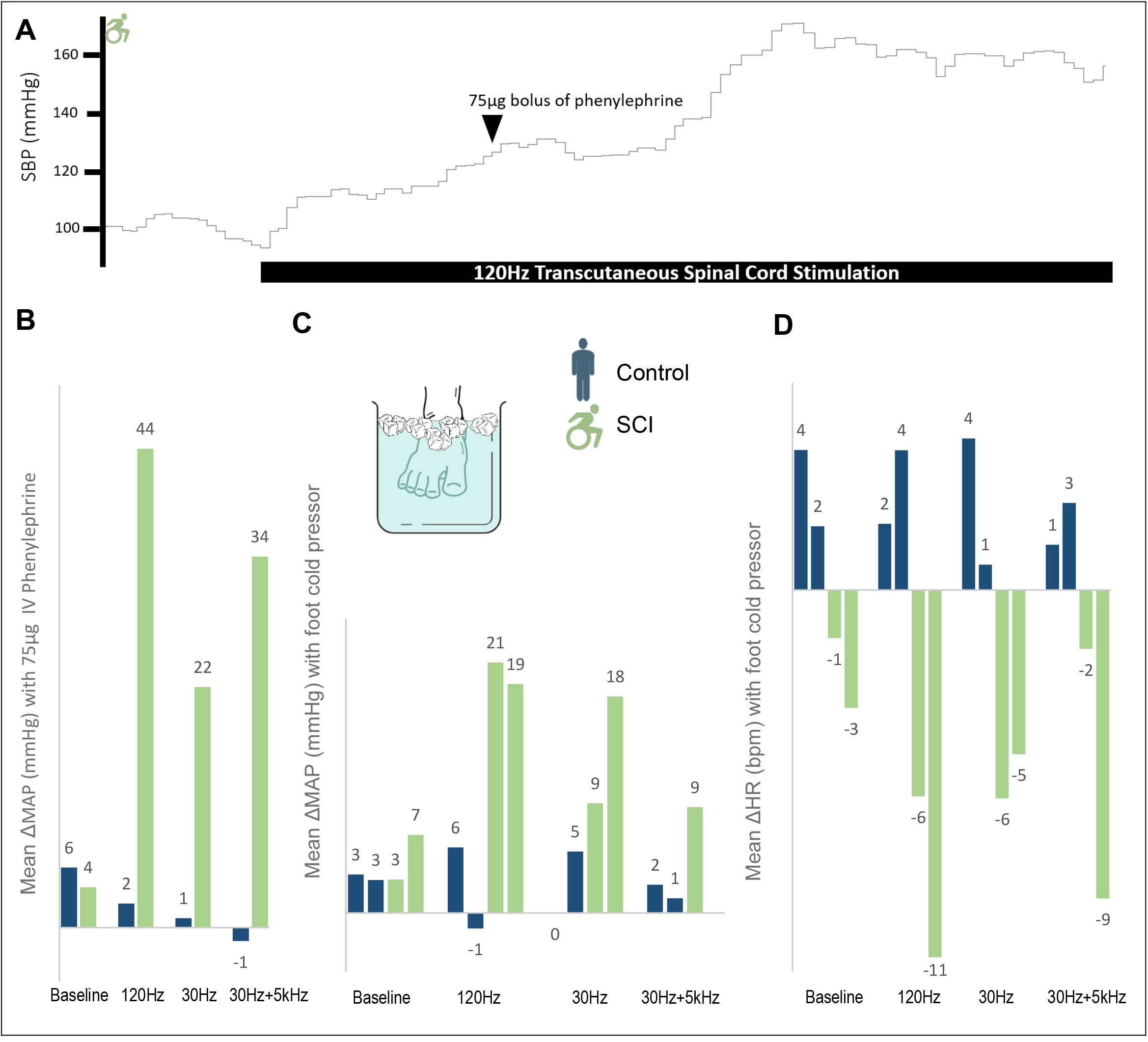
**A)** Representational trace of SBP with bolus phenylephrine and transcutaneous spinal cord stimulation. **B)** MAP change with 75μg dose of intravenous phenylephrine per frequency for matched pair 1 (uninjured control and individual with SCI). Change was calculated as 30 seconds of stable baseline recordings immediately prior to each successive dose compared to a 20 second window centered at initial nadir in HR, when maximal effect was identified. The addition of stimulation caused significantly greater ΔMAP in SCI compared to control (33.5±6.4mmHg vs 0.6±1.0mmHg, p=0.03). **C, D)** MAP and HR changes with foot cold pressor test. *Of note, missing 30Hz+5kHz MAP due to equipment malfunction for second individual with SCI*. While controls exhibit the expected tachycardic response with foot cold pressor, individuals with SCI conversely have bradycardia, indicating a disengaged baroreflex pathway attempting to compensate for sympathetic tone generated below the level of injury. Analyses compared 3 minutes of stable baseline with 20 second window around the peak MAP. Abbreviations. SBP= Systolic Blood Pressure, MAP= Mean Arterial Pressure, HR= Heart Rate

## Discussion

Transcutaneous spinal cord stimulation consistently caused tonic sympathetic activation in a frequency-dependent manner after SCI and had no effect in matched, uninjured controls. Tonic sympathetic activation from stimulation agrees with previous results demonstrating restored blood pressure maintenance during head-up-tilt when sympathoexcitation is needed. Our results demonstrate that this tonic sympathetic activation can further restore phase II of Valsalva’s maneuver. However, stimulation has no beneficial effect on sympathoinhibitory responses and may promote AD responses to noxious stimuli below the level of lesion.

This last effect is of significant clinical importance. For example, during 120Hz stimulation with foot cold pressor, one individual with SCI demonstrated an almost 20mmHg rise in MAP despite an 11-beat bradycardia; this indicates that there was massive vasoconstriction below the level of lesion in response to the cold noxious stimulus and an ongoing decoupled HR response – opposite of what would be expected with improved autonomic regulation.

Taken together, these tests demonstrate that spinal cord stimulation did not improve dynamic autonomic regulation. Further, AD is a potentially dangerous condition and future studies should adequately monitor for AD with stimulation, since individuals are often asymptomatic.^8^ This is the first head-to-head study comparing previously advocated frequencies of spinal cord stimulation in a comprehensive manner.

Stimulation at 120Hz had the greatest degree of sympathetic activation, followed by 30Hz+5kHz, and then 30Hz. This effect was mirrored in all study tests.

## Limitations

While this sample size is typical of the growing field^4-7^, generalizability may be limited with a small cohort. Despite this, across 92 independent autonomic tests to systematically characterize autonomic deficits and stimulation at different frequencies, results were remarkably consistent. Most especially, the previously undocumented risk of AD warrants notice.

## Conclusions

Transcutaneous spinal cord stimulation causes tonic sympathetic activation in a frequency-dependent manner, without evidence of improvements in autonomic regulation after SCI. This may lower the threshold for autonomic dysreflexia.

## Data Availability

Data produced in the present study will be made available upon publication with link activated in Harvard Dataverse.

---

**eFigure S1:**
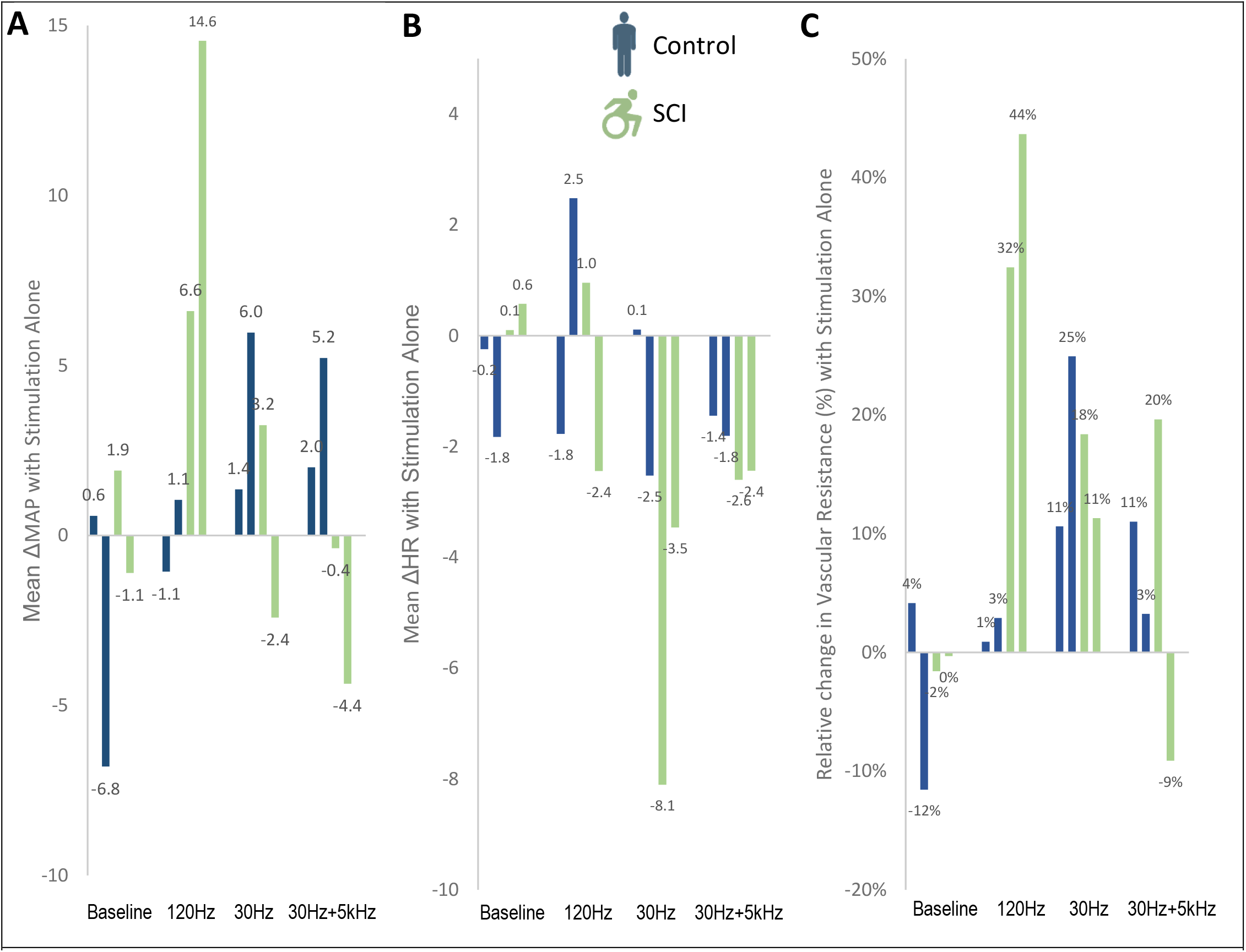
Physiologic response to transcutaneous spinal cord stimulation alone in a resting state **A)** Mean change in MAP with stimulation alone. **B)** Mean changes in HR with stimulation alone. **C)** Relative changes in lower extremity vascular resistance with stimulation alone. Grouping stimulation frequencies, vascular resistance increased an average of 19.0±7.7% for individuals with SCI, vs 8.9±3.6% for controls (p=0.32). The greatest effect was seen at 120Hz for those with SCI, with resting vascular resistance increasing 32.4% and 43.7% respectively. At this same stimulation frequency in matched controls, vascular resistance increased 0.9% and 2.9%. All other stimulation frequencies had minimal impact in this unchallenged state. Abbreviations. MAP= Mean Arterial Pressure, HR= Heart Rate

## Acknowledgements

Research reported in this publication was supported by pilot funding from the National Institutes of Health National Center of Neuromodulation for Rehabilitation, the National Center for Complementary and Integrative Health, the National Institute on Deafness and Other Communication Disorders, and the National Institute of Neurological Disorders and Stroke. NIH/NICHD Grant Number P2CHD086844 which was awarded to the Medical University of South Carolina. The contents are solely the responsibility of the authors and do not necessarily represent the official views of the NIH or NICHD. Dr. Solinsky’s time was protected by K23HD102663 through NIH/NICHD.

## References

1. Wecht JM, Bauman WA. Implication of altered autonomic control for orthostatic tolerance in SCI. Auton Neurosci. 2018;209:51–58.

2. Solinsky R, Kirshblum SC, Burns SP. Exploring detailed characteristics of autonomic dysreflexia. J Spinal Cord Med. 2018;41(5):549–555.

3. Teasell RW, Arnold JMO, Krassioukov A, Delaney GA. Cardiovascular consequences of loss of supraspinal control of the sympathetic nervous system after spinal cord injury. Arch Phys Med Rehabil. 2000;81:506–516.

4. Squair JW, Gautier M, Mahe L, et al. Neuroprosthetic baroreflex controls haemodynamics after spinal cord injury. Nature. 2021;590(7845):308–314.

5. West CR, Phillips AA, Squair JW, et al. Association of Epidural Stimulation with Cardiovasular Function in an Individual With Spinal Cord Injury. JAMA Neurol. 2018;75:630–632.

6. Harkema SJ, Wang S, Angeli CA, et al. Normalization of blood pressure with spinal cord epidural stimulation after severe spinal cord injury. Front Hum Neurosci. 2018;12(83):1–11.

7. Kaur B, Arumugam N. Effect of surface spinal stimulation on autonomic nervous system in the patients with spinal cord injury. Arch Med Health Sci. 2014;2(2):126–130.

8. Burns M, Solinsky R. Toward rebalancing blood pressure instability after spinal cord injury with spinal cord electrical stimulation: A mini review and critique of the evolving literature. Auton Neurosci. 2022;237:102905

9. Burns SP, Tansey KE. The expedited international standards for neurological classification of spinal cord injury (E-ISNCSCI). Spinal Cord. 2020;58(6):633–634.

10. Nishimura, RA, Tajik AJ. The Valsalva maneuver and response revisited. Mayo Clin Proc. 1986;61(3):211–217.

11. La Rovere MT, Specchia G, Mortara A, Schwartz PJ. Baroreflex sensitivity, clinical correlates, and cardiovascular mortality among patients with a first myocardial infarction. A prospective study. Circulation. 1988;78(4):816–824.

12. Hunt BE, Fahy L, Farquhar WB, Taylor JA. Quantification of mechanical and neural components of vagal baroreflex in humans. Hypertension. 2001;37(6):1362–1368.

13. Victor RG, Leimbach Jr WN, Seals DR, Wallin BG, Mark AL. Effects of the cold pressor test on muscle sympathetic nerve activity in humans. Hypertension. 1987;9(5):429–436.

14. Vogel ER, Sandroni P, Low PA. Blood pressure recovery from Valsalva maneuver in patients with autonomic failure. Neurology. 2005;65(10):1533–1537.

